## Supplementary material for "Applying behavioural economics principles to increase demand for free HIV testing services at private doctor-led clinics in Johannesburg, South Africa: A randomised controlled trial"

**Supplementary table 1:** Proportion of individuals presenting at the GP practice over the total number of brochures distributed by study arm

**Supplementary Table 2.** Sensitivity analysis of Odds ratios (95% CI) from exploratory logistic regression results comparing SOC, healthy lifestyle brochure and care recipient voucher brochure arms

**Supplementary Table 3:** Effect of intervention on odds of presenting at the GP practice with sociodemographics

**Supplementary Table 1:** Proportion of individuals presenting at the GP practice over the total number of brochures distributed by study arm

| | | Standard of Care | | Healthy Lifestyle Brochure | | Recipient Care Voucher | |
|---|---|---|---|---|---|---|---|
| | | **Brochures distributed** | **Individuals presenting** | **Brochures distributed** | **Individuals presenting** | **Brochures distributed** | **Individuals presenting** |
| | | n | n (%) | n | n (%) | n | n (%) |
| **Total** | | 3802 | 137 (3.6) | 3840 | 153 (4.0) | 3829 | 158 (4.1) |
| **Gender** | Male | 2246 | 69 (3.1) | 2293 | 76 (3.3) | 2235 | 82 (3.7) |
| | Female | 1532 | 68 (4.4) | 1512 | 77 (5.1) | 1557 | 76 (4.9) |
| **Age group** | 18-24 years | 654 | 24 (3.7) | 612 | 17 (2.8) | 624 | 24 (3.8) |
| | 25-34 years | 1901 | 67 (3.5) | 1911 | 60 (3.1) | 1962 | 88 (4.5) |
| | 35-44 years | 1015 | 37 (3.6) | 1060 | 48 (4.5) | 991 | 35 (3.5) |
| | ≥ 45 years | 216 | 9 (4.2) | 228 | 28 (12.3) | 225 | 11 (4.9) |
| **HIV test outcomes** | HIV negative | - | 130 (3.4) | - | 146 (3.8) | - | 156 (4.1) |
| | PLHIV | - | 4/137 (0.1) | - | 5/153 (0.1) | - | 1/158 (0.0) |
| | Unknown HIV outcomes | - | 3 (0.1) | - | 2 (0.1) | - | 1 (0.0) |

**Supplementary Table 2:** Sensitivity analysis of Odds ratios (95% CI) from exploratory logistic regression results comparing SOC, healthy lifestyle brochure and care recipient voucher brochure arms*

| | | No. of participants | Presenting at the GP practice (%) | UOR (95% CI) | P-value | AOR (95% CI)** | P-value |
|---|---|---|---|---|---|---|---|
| **Study arm** | SOC | 3603 | 137 (3.8%) | 1 [Ref] | | 1 [Ref] | |
| | HLS | 3620 | 153 (4.2%) | 1.11 (0.88-1.40) | .394 | 0.92 (0.70-1.21) | .554 |
| | RCV | 3608 | 158 (4.4%) | 1.12 (0.89-1.41) | .347 | 0.99 (0.76-1.23) | .924 |
| **Gender** | Male | 6406 | 227 (3.5%) | 1 [Ref] | | 1 [Ref] | |
| | Female | 4298 | 221 (5.1%) | 1.11 (0.89-1.38) | .371 | 0.97 (0.77-1.21) | .762 |
| **Age group** | 18-24 | 1744 | 65 (3.7%) | 1 [Ref] | | 1 [Ref] | |
| | 25-34 | 5438 | 215 (4.0%) | 0.76 (0.56-1.01) | .061 | 0.95 (0.70-1.28) | .728 |
| | 35-44 | 2927 | 120 (4.1%) | 0.76 (0.55-1.05) | .101 | 0.89 (0.64-1.63) | .509 |
| | ≥ 45 | 612 | 48 (7.8%) | 1.31 (0.84-2.04) | .230 | 1.04 (0.66-1.63) | .865 |
| **GP group** | GP group with limited visibility | 6630 | 44 (0.7) | 1 [Ref] | | 1 [Ref] | |
| | GP group with high visibility | 4201 | 404 (9.6) | 6.49 (5.15-8.18) | .000 | 4.92 (3.82-6.34) | .000 |
| **Language** | English | 7756 | - | 1 [Ref] | | 1 [Ref] | |
| | IsiZulu | 2466 | - | 1.50 (1.21-1.85) | .000 | 1.06 (0.82-1.37) | .665 |
| | SeSotho | 609 | - | 1.57 (1.09-2.26) | .016 | 0.63 (0.33-1.20) | .163 |

* data excludes all duplicates

**Adjusted for study arm category, gender, age group, GP group and language

**Supplementary Table 3.** Effect of intervention on odds of presenting at the GP practice with sociodemographics*

| | | No. of participants | Presenting at the GP practice (%) | UOR (95% CI) | P-value | AOR 95% (CI) | P-value |
|---|---|---|---|---|---|---|---|
| **Study arm** | SOC | 3802 | 137 (3.6%) | 1 [Ref] | | 1 [Ref] | |
| | HLS | 3840 | 153 (4.0%) | 1.11 (0.88-1.40) | .384 | 1.02 (0.79-1.32) | .873 |
| | RCV | 3829 | 158 (4.1%) | 1.15 (0.91-1.45) | .236 | 1.08 (0.84-1.40) | .559 |
| **Gender** | Male | 6774 | 227 (3.4%) | 1 [Ref] | | 1 [Ref] | |
| | Female | 4601 | 221 (4.8%) | 1.11 (0.89-1.38) | .371 | 1.02 (0.82-1.26) | .852 |
| **Age group** | 18-24 | 1890 | 65 (3.4%) | 1 [Ref] | | 1 [Ref] | |
| | 25-34 | 5774 | 215 (3.7%) | 0.82 (0.62-1.08) | .155 | 1.03 (0.77-1.37) | .856 |
| | 35-44 | 3066 | 120 (3.9%) | 0.77 (0.56-1.06) | .108 | 0.90 (0.65-1.24) | .505 |
| | ≥ 45 | 669 | 48 (7.2%) | 1.27 (0.83-1.94) | .274 | 0.98 (0.63-1.51) | .918 |
| **GP group** | GP group with limited visibility | 6885 | 44 (0.6%) | 1 [Ref] | | 1 [Ref] | |
| | GP group with high visibility | 4586 | 404 (8.8%) | 6.13 (4.86-7.72) | .000 | 5.30 (4.14-6.79) | .000 |
| **Language**** | English | 8165 | - | 1 [Ref] | | 1 [Ref] | |
| | IsiZulu | 2656 | - | 1.46 (1.18-1.80) | .000 | 0.98 (0.77-1.26) | .895 |
| | SeSotho | 650 | - | 1.49 (1.03-2.16) | .035 | 1.07 (0.67-1.72) | .762 |

*Adjusted for gender, age group, clinic group, language (using combined GP groups and combined intervention arms)

**Language was not recorded at presentation at the GP practice
